# Seroincidence of SARS-CoV-2 infection prior to and during the rollout of vaccines in a community-based prospective cohort of U.S. adults

**DOI:** 10.1101/2023.09.29.23296142

**Authors:** Denis Nash, Avantika Srivastava, Jenny Shen, Kate Penrose, Sarah Gorrell Kulkarni, Rebecca Zimba, William You, Amanda Berry, Chloe Mirzayi, Andrew Maroko, Angela M. Parcesepe, Christian Grov, McKaylee M. Robertson

**Affiliations:** Institute for Implementation Science in Population Health (ISPH), City University of New York (CUNY); New York, New York, USA; Department of Epidemiology and Biostatistics, Graduate School of Public Health and Health Policy, City University of New York (CUNY); New York, New York, USA; Department of Environmental, Occupational, and Geospatial Health Sciences, Graduate School of Public Health and Health Policy, City University of New York (CUNY); New York, New York, USA; Department of Maternal and Child Health, Gillings School of Public Health, University of North Carolina, Chapel Hill, North Carolina, USA; Carolina Population Center, University of North Carolina at Chapel Hill, Chapel Hill, North Carolina, USA; Department of Community Health and Social Sciences, Graduate School of Public Health and Health Policy, City University of New York (CUNY); New York, New York, USA

**Keywords:** COVID-19, serology, infection-induced seroconversion, asymptomatic infection, physical distancing, natural history study, epidemiologic study, essential workers, public health interventions, community transmission

## Abstract

**LONG ABSTRACT:** *Background:* Infectious disease surveillance systems, which largely rely on diagnosed cases, underestimate the true incidence of SARS-CoV-2 infection, due to under-ascertainment and underreporting. We used repeat serologic testing to measure N-protein seroconversion in a well-characterized cohort of U.S. adults with no serologic evidence of SARS-CoV-2 infection to estimate the incidence of SARS-CoV-2 infection and characterize risk factors, with comparisons before and after the start of the SARS-CoV-2 vaccine and variant eras.

*Methods:* We assessed the incidence rate of infection and risk factors in two sub-groups (cohorts) that were SARS-CoV-2 N-protein seronegative at the start of each follow-up period: 1) the pre-vaccine/wild-type era cohort (n=3,421), followed from April to November 2020; and 2) the vaccine/variant era cohort (n=2,735), followed from November 2020 to June 2022. Both cohorts underwent repeat serologic testing with an assay for antibodies to the SARS-CoV-2 N protein (Bio-Rad Platelia SARS-CoV-2 total Ab). We estimated crude incidence and sociodemographic/epidemiologic risk factors in both cohorts. We used multivariate Poisson models to compare the risk of SARS-CoV-2 infection in the pre-vaccine/wild-type era cohort (referent group) to that in the vaccine/variant era cohort, within strata of vaccination status and epidemiologic risk factors (essential worker status, child in the household, case in the household, social distancing).

*Findings:* In the pre-vaccine/wild-type era cohort, only 18 of the 3,421 participants (0.53%) had ≥1 vaccine dose by the end of follow-up, compared with 2,497/2,735 (91.3%) in the vaccine/variant era cohort. We observed 323 and 815 seroconversions in the pre-vaccine/wild-type era and the vaccine/variant era and cohorts, respectively, with corresponding incidence rates of 9.6 (95% CI: 8.3-11.5) and 25.7 (95% CI: 24.2-27.3) per 100 person-years. Associations of sociodemographic and epidemiologic risk factors with SARS-CoV-2 incidence were largely similar in the pre-vaccine/wild-type and vaccine/variant era cohorts. However, some new epidemiologic risk factors emerged in the vaccine/variant era cohort, including having a child in the household, and never wearing a mask while using public transit. Adjusted incidence rate ratios (aIRR), with the entire pre-vaccine/wild-type era cohort as the referent group, showed markedly higher incidence in the vaccine/variant era cohort, but with more vaccine doses associated with lower incidence: aIRR_un/undervaccinated_=5.3 (95% CI: 4.2-6.7); aIRR_primary series only_=5.1 (95% CI: 4.2-7.3); aIRR_boosted once_=2.5 (95% CI: 2.1-3.0), and aIRR_boosted twice_=1.65 (95% CI: 1.3-2.1). These associations were essentially unchanged in risk factor-stratified models.

*Interpretation:* In SARS-CoV-2 N protein seronegative individuals, large increases in incidence and newly emerging epidemiologic risk factors in the vaccine/variant era likely resulted from multiple co-occurring factors, including policy changes, behavior changes, surges in transmission, and changes in SARS-CoV-2 variant properties. While SARS-CoV-2 incidence increased markedly in most groups in the vaccine/variant era, being up to date on vaccines and the use of non-pharmaceutical interventions (NPIs), such as masking and social distancing, remained reliable strategies to mitigate the risk of SARS-CoV-2 infection, even through major surges due to immune evasive variants. Repeat serologic testing in cohort studies is a useful and complementary strategy to characterize SARS-CoV-2 incidence and risk factors.

**SHORT ABSTRACT:** This study used repeat serologic testing to estimate infection rates and risk factors in two overlapping cohorts of SARS-CoV-2 N protein seronegative U.S. adults. One mostly unvaccinated sub-cohort was tracked from April to November 2020 (pre-vaccine/wild-type era, n=3,421), and the other, mostly vaccinated cohort, from November 2020 to June 2022 (vaccine/variant era, n=2,735). Vaccine uptake was from 0.53% and 91.3% in the pre-vaccine and vaccine/variant cohorts, respectively. Corresponding seroconversion rates were 9.6 and 25.7 per 100 person-years. In both cohorts, sociodemographic and epidemiologic risk factors for infection were similar, though new risks emerged in the vaccine/variant era, such as having a child in the household. Despite higher incidence rates in the vaccine/variant cohort, vaccine boosters, masking, and distancing likely reduced infection risk, even through major variant surges. Repeat serologic testing in cohorts is a useful and complementary strategy to characterize incidence and risk factors.

*Funding:* The work was supported by the CUNY Institute for Implementation Science in Population Health, the U.S. National Institutes of Allergy and Infectious Diseases (NIAID), Pfizer, Inc., and the U.S. National Institute of Mental Health (NIMH).

## INTRODUCTION

Infectious disease surveillance systems, which largely rely on diagnosed case counts, emergency department visits, hospital admissions, and deaths, underestimate the true incidence of infection due to asymptomatic infections, under-ascertainment/diagnosis, and underreporting to health departments. This has proved to be a challenge during the COVID-19 pandemic in the United States (US)^1,2^, where the national surveillance system relied on the reporting of positive SARS-CoV-2 real-time reverse-transcription polymerase chain reaction (RT-PCR) test results by providers and laboratories. These data have effectively been used as a proxy for the incidence and prevalence of SARS-CoV-2 infection in the US, including to inform pandemic policy decisions, often with no efforts to adjust for underestimation due to asymptomatic infections, testing and healthcare access, evolving testing practices, behaviors, and policies.^2–5^ Studies have shown that estimates of infection based on seroprevalence far exceed the number of diagnosed and reported cases.^2,6–9^ These issues have posed major challenges to using surveillance data to assess the true burden of infection and related risk factors and inform decision-making in an evolving pandemic^10^ These issues are magnified with the end of the national emergency declaration in May 2023, as surveillance for SARS-CoV-2 further dismantles, testing behaviors change and official case counts continue to fall.^11^

The risk of SARS-CoV-2 infection is determined by multiple factors, including frequency of exposure, underlying medical conditions, vaccination status, virus properties and their evolution, levels of community transmission, and individual behaviors, such as the use of non-pharmaceutical interventions (NPIs) like masking and social distancing.^12–15^ Vaccines, which were authorized by the FDA on December 11, 2020 and started to become more widely available in the United States in March 2021^16^, have dramatically reduced the risk of severe disease and death from SARS-CoV-2^17^, including during the Alpha (March-June 2021), Delta (June-December 2021) and Omicron (December 2021-present) variant eras.^18^ However, with the emergence of the Delta variant^19^ and particularly during the Omicron era, vaccine effectiveness *against infection* reduced and waned quickly after vaccination, including after a booster.^18,20–23,15–18^ Recent population-representative, cross-sectional studies during successive Omicron surges have shown a high point prevalence of SARS-CoV-2 infection among those who were previously vaccinated and boosted.^2,24^

Alongside variant surges, starting in late 2021, the United States relaxed public health policies and guidelines recommending or requiring quarantine and isolation, masking, social distancing, and remote K-12 schooling in response to the increasing availability and uptake of vaccines and the availability of effective therapeutics.^25–29^ National policies have increasingly emphasized the importance of using vaccines and boosters to reduce the burden of severe disease and healthcare system strain, with less emphasis on NPIs for preventing infection.^14^ These policy choices and related public health messaging likely impacted individual-level and community-level risk factors and behavior, resulting in less utilization of NPIs such as masking, social distancing, and more. ^30,31^

Beyond basic demographics and geography, risk factors for SARS-CoV-2 infection (vs. diagnoses), including asymptomatic infection, have not been well-characterized in the vaccine and variant eras, either via routine case-based surveillance or by cross-sectional seroprevalence studies in population-based samples.^10,32,33^ Moreover, the degree of the potential protective effect of vaccines for preventing SARS-CoV-2 infection has not been examined in a population-based prospective cohort with systematically gathered, time-updated information on risk factors, behaviors, and infection status derived from infection-induced seroconversions.

Within a well-characterized national prospective cohort of US adults (for whom we previously characterized risk factors for seroincident SARS-CoV-2 infection in the pre-vaccine/wild-type era^15^), we used repeat serologic testing to assess the incidence of SARS-CoV-2 infection and risk factors among those who were N protein seronegative by March 2021 (the start of the variant era), with vaccine uptake and infection status assessed through June 2022.

## METHODS

### Recruitment

We used internet-based strategies^34–36^ to recruit a geographically and socio-demographically diverse cohort of 6,740 adults into longitudinal follow-up with at-home, dried blood spot (DBS) specimen collection. To be eligible for inclusion in the cohort, individuals had to: 1) reside in the United States or a U.S. territory; 2) be ≥18 years of age; 3) provide a valid email address for follow-up; and 4) demonstrate early engagement in study activities (provision of a baseline specimen for serologic testing or completion of >1 recruitment/enrollment visit). Details of the study design and recruitment procedures^36^ and a pre-vaccine/wild-type era serology-based incidence study in this cohort^15^ are described elsewhere. Cohort enrollment was completed between March 28 and August 21, 2020, during which baseline demographic data collection took place. The full cohort includes participants from all 50 U.S. states, the District of Columbia, Puerto Rico, and Guam.

### Study population

The study population (n=3,582) was divided into two *overlapping* sub-groups (henceforth cohorts, Figure 1) which broadly correspond to those with a seronegative specimen during Serology Period 1 with at least one follow-up specimen (pre-vaccine/wild-type era cohort, n=3,421), and those with a seronegative specimen during Serology Period 2 with at least one follow-up specimen (vaccine/variant era cohort, n=2,735) (Figure 2).^37,38^ The vaccine/variant era cohort included those in the pre-vaccine/wild-type era cohort who remained seronegative on their specimen from Serology Period 2 (Figure 2).

**Figure 1.**
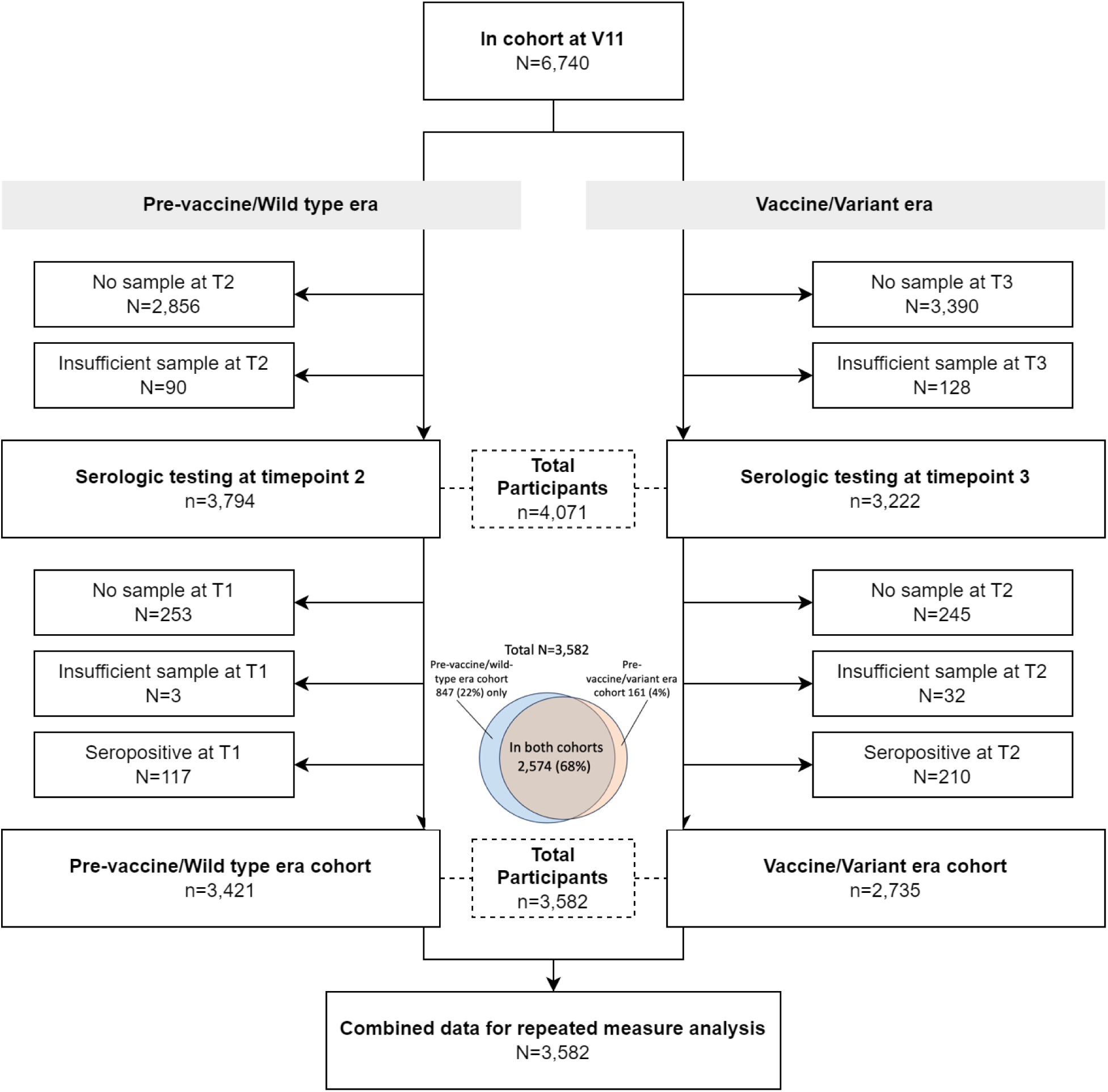
Study population and sub-cohorts

**Figure 2.**
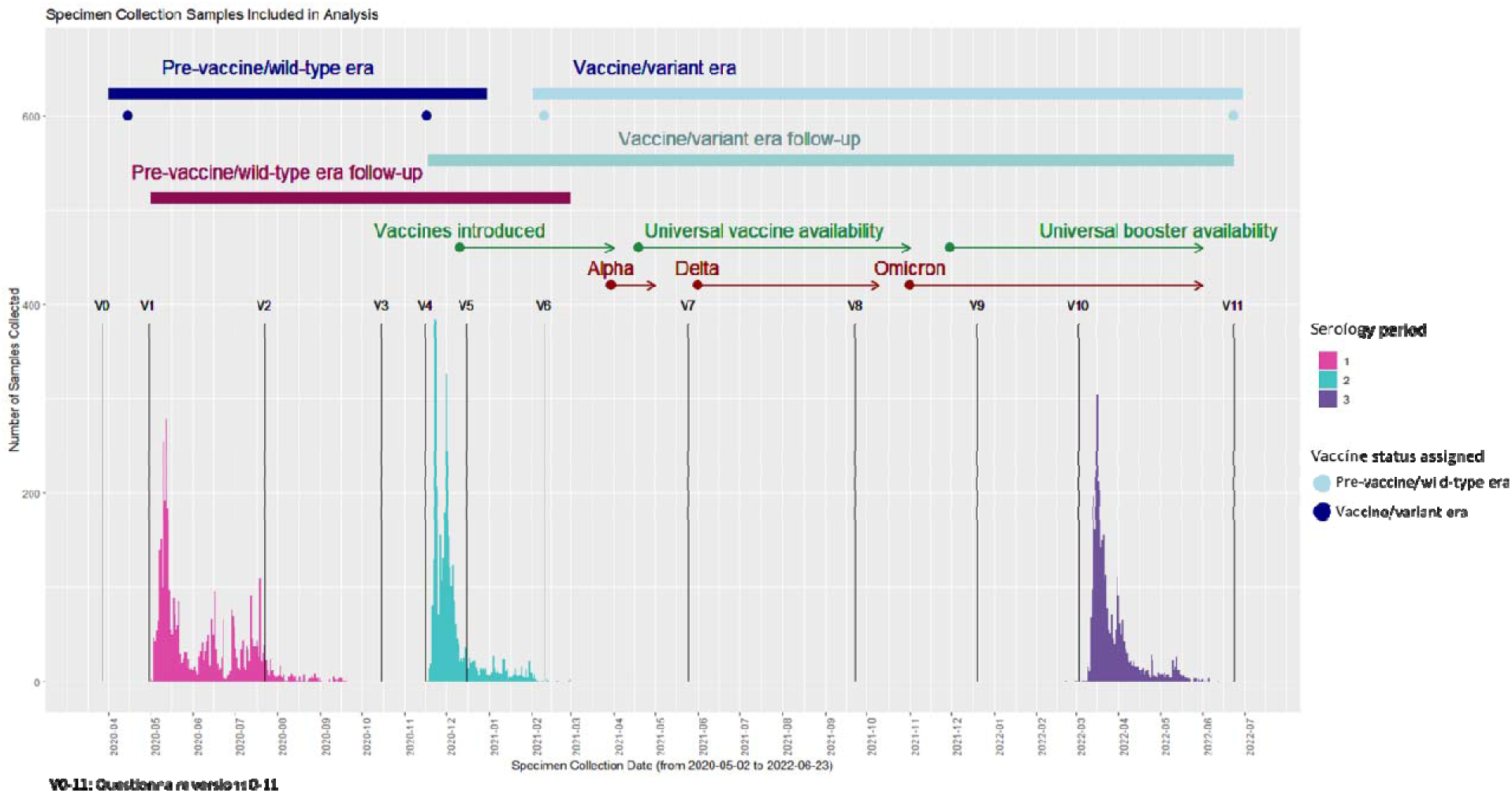
Timing of specimen collection, vaccine rollout, and cohort follow-up

### Follow-up data collection

From 14 follow-up study encounters occurring approximately quarterly between August 2020 and July 2022, we obtained repeated measurements of epidemiologic risk factors, COVID-19 symptoms, non-study-related SARS-CoV-2 testing (PCR or rapid, at-home rapid), hospitalizations, use of NPIs, public health strategies (i.e., quarantine, isolation), and contact tracing encounters.

### Serologic testing

Figure 2 shows the three periods of serologic testing in the cohort – from April through September 2020 (Serology Period 1), November 2020 through March 2021 (Serology Period 2), and March 2022 through June 2022 (Serology Period 3). During these periods, participants were invited to complete serologic testing using an at-home self-collected dried blood spot (DBS) specimen collection kit. DBS cards were sent from and returned to the study laboratory (Molecular Testing Laboratories [MTL], Vancouver, WA) via the U.S. Postal Service using a self-addressed, stamped envelope containing a biohazard bag.

To assess infection-induced seroconversion, all DBS specimens were tested by the study laboratory for total antibodies to the SARS-CoV-2 nucleocapsid protein (total nucleocapsid Ab) using the Bio-Rad Platelia test for IgA, IgM, and IgG (manufacturer sensitivity 98.0%, specificity 99.3%).^39^ Other studies have independently validated this assay and found average sensitivity and specificity of 91.7% and 98.8%, respectively.^40–42^ This assay was also validated for use with DBS by the study laboratory, which found 100% sensitivity and 100% specificity (MTL, personal communication).

### Outcome (infection-induced seroconversion)

Among those individuals with two total nucleocapsid Ab tests, the outcome of infection-induced SARS-CoV-2 seroconversion was defined as having a negative total nucleocapsid Ab test in Serology Period 1 followed by a positive total nucleocapsid Ab test in Serology Period 2 (within the pre-vaccine/wild-type era cohort) or a negative total nucleocapsid Ab test in Serology Period 2 followed by a positive total nucleocapsid Ab test in Serology Period 3 (within the vaccine/variant era cohort). We estimated person-years of follow-up in each cohort using the collection dates for each specimen. Participants could contribute person-time to each cohort (pre-vaccine/wild-type or vaccine/variant era). When the specimen collection date was missing, we used the date the laboratory received the sample. For those who seroconverted, the seroconversion date was assigned as the midpoint between the initial seronegative specimen collection date and earlier of: 1) the date of the follow-up seropositive specimen collection; or 2) the reported date of a positive SARS-CoV-2 test result (PCR or rapid test) in between the initial and follow-up specimen collection dates.

### Exposures

#### Timing of exposure data collection on risk factors, behaviors, and vaccination status

All exposure measurements were derived from time-updated questionnaire data collected during each era, and only those measures taken prior to outcome measurement for a given era were used. For the pre-vaccine/wild-type era cohort, we used exposure data from the questionnaire for study visits 1 through 4 (V1-V4; Figure 2). For the vaccine/variant era cohort, we used data from V4-V10 questionnaires (Figure 2).

#### Individual-Level COVID-19 Risk Factors

We collected time-updated information on an array of epidemiologic risk factors for SARS-CoV-2 infection reported by participants, including the following: essential worker status (those working in healthcare, emergency response, law enforcement, delivery of food/goods, transportation), household factors (household crowding defined as ≥4 people living in a single unit of a multi-unit dwelling, having a child in the household, and having a confirmed COVID-19 case in a household member before participant tested positive); spending time in public places (attending mass gatherings, indoor dining in a restaurant or bar, outdoor dining at a restaurant or bar, visiting places of worship, or visiting public parks or pools); mask use indoors (for grocery shopping, visiting non–household members, at work, and in salons or gyms); mask use outdoors; gathering in groups with 10 or more people; travel during the pandemic (air travel and public transit use); and individual-level factors that may increase the risk of infection and/or severe COVID-19 (comorbid conditions, binge drinking, regular cannabis use or un-prescribed opioid use). Binge drinking was defined as six or more drinks in one sitting during the last month, asked as part of the Alcohol Use Disorders Identification Test questions on select questionnaires.^43^ As a measure of susceptibility to severe COVID-19, we used comorbid conditions or exposures that CDC identified as increasing the risk for COVID-19 complications, given SARS-CoV-2 infection: age >=60 years, daily smoking, chronic lung disease, including chronic obstructive pulmonary disease, emphysema, chronic bronchitis, serious heart conditions, current asthma, type 2 diabetes, kidney disease, immunocompromised condition, or an HIV diagnosis.^44^

#### Risk groups

We hypothesized that some participants may be at higher risk of SARS-CoV-2 infection in the vaccine/variant era because of membership in a group more directly affected by policy changes, including changes to guidelines and public health messaging. These groups included essential workers, those living in crowded households, and those with children in the household who might attend childcare or school. For essential worker status, household factors, and other binary variables, we assigned exposure status based on any vs. no exposure (e.g., having a confirmed COVID-19 case in the household or not) that occurred within each era.

#### Risk behaviors

We also hypothesized that some participants may have a higher risk of SARS-CoV-2 infection in the vaccine/variant era relative to the pre-vaccine/wild-type era due to the de-implementation of policies that may change risk factors and behaviors. These risk factors/behaviors included: mask use indoors while visiting non-household members, mask use at work, social distancing with individuals the participant knows, and social distancing with individuals the participant does not know. For time-dependent exposure variables (e.g., social distancing and masking), we assigned exposure status based on a hierarchy of exposure risk during follow-up. Specifically, participants were classified according to the highest risk strata (e.g., never masking>sometimes masking>always masking) that they reported at one or more follow-up assessments.

#### Composite risk score

We computed a composite COVID-19 risk score, as many of the above COVID-19 risk groups and behaviors are likely to be highly correlated. We applied least absolute shrinkage selection operator (LASSO) regression to select the set of risk factors that best-predicted seroconversion in the pre-vaccine/wild-type era.^45^ The LASSO model selected household crowding, having a confirmed COVID-19 case in a household member, indoor dining in a bar/restaurant, gathering with groups of ≥10, and no mask use indoors in salons or gyms as the most predictive of seroconversion in our cohort during the pre-vaccine/wild-type era. Scores were assigned to each participant based on their responses for each of the risk factors selected by the LASSO model. Sores were normalized between 0 and 100, with higher scores indicating more engagement in high-risk activities (details in Statistical Appendix). The composite score was divided into tertiles for analysis.

#### Vaccination status

For the pre-vaccine/wild-type cohort, vaccination status *at the start of follow-up* was assigned as unvaccinated for all participants in the cohort, since vaccines were not available during Serology Period 1 (i.e., 100% were unvaccinated at the start). Vaccination status *at the end of follow-up* was assigned at the start of Serology Period 2 (based on responses to the V4 questionnaire in Figure 2), at which time almost the entire cohort (99%) remained unvaccinated. For the vaccine/variant era cohort, vaccination status *at the start of follow-up* was assigned based on vaccination status as of February 10, 2021, which corresponded with the first questionnaire after Serology Period 2 specimen collection (V6 in Figure 2). Individuals in the vaccine/variant-era cohort were classified according to their vaccination/booster status *as of the end of follow-up* (June 2022), with categories as follows: un/undervaccinated (unvaccinated or did not complete primary vaccine series), completed primary vaccine series, completed primary vaccine series with one booster, and completed primary vaccine series with two or more boosters. Completing a primary vaccine series was defined as one dose for participants who indicated that they had received the Johnson & Johnson vaccine and two doses for any other COVID vaccine type specified.

### Statistical analysis

Seroincidence of SARS-CoV-2 infection was calculated within each cohort and across strata of sociodemographic factors and epidemiologic risk factors, selected based on the literature and on previous published pre-vaccine/wild-type era analyses of SARS-CoV-2 incidence in this cohort.^15^ Crude associations of each factor with SARS-CoV-2 infection were reported as rate ratios. A multivariable mixed effects Poisson model with random coefficients, using the log of total person-time as the offset and an unstructured covariance matrix, was used to estimate the rate ratio of incident SARS-CoV-2 infection stratified by vaccination status (un/undervaccinated, vaccinated, boosted once, boosted more than once) in the vaccine/variant era cohort. The entire pre-vaccine/wild-type era cohort was used as the referent group in these models. We ran a crude and multivariable overall model. To assess the association of vaccination status within different risk factor strata, we ran 12 multivariable models, one for each stratum of five different risk factor groups [essential workers, household children, household cases, social distancing with those you know, and social distancing with those you don’t know]. All multivariable models were adjusted for age, sex, and the presence of comorbidities. All mixed models accounted for repeated measures among participants by including a random intercept for subject. All data were cleaned and analyzed in R and SAS.

### Ethical Approval

The study protocol was approved by the Institutional Review Board at the City University of New York (CUNY).

## RESULTS

### Sample characteristics

The characteristics of the study cohorts are shown in Table 1. Seventy-two percent of subjects (n=2,574) were represented in both cohorts, 24% (n=847) were represented only in the pre-vaccine/wild-type era cohort and 4% (n=161) were represented only in the vaccine/variant era cohort (Figure 1). Participants in each cohort were very similar on measured characteristics, except for employment status, where a slightly lower proportion was unemployed in the vaccine/variant era than in the pre-vaccine/wild-type era (6.5% vs 11.1%, respectively).

**Table 1.** Characteristics of study participants in each cohort.

|  | Pre-vaccine/wild-type era |  | Vaccine/variant era |  |
| --- | --- | --- | --- | --- |
|  | N | % | N | % |
| **Total** | 3421 | 100 | 2735 | 100 |
| **No. participants included in both cohorts** | 2574 | 75.24 | 2574 | 94.11 |
| **No. participants included one cohort** | 847 | 24.76 | 161 | 5.89 |
| **Age** |  |  |  |  |
| Median (IQR) | 42 (32-56) |  | 42 (33, 57) |  |
| 18-29 | 617 | 18.04 | 457 | 16.71 |
| 30-39 | 934 | 27.3 | 742 | 27.13 |
| 40-49 | 654 | 19.12 | 518 | 18.94 |
| 50-59 | 514 | 15.02 | 428 | 15.65 |
| 60+ | 702 | 20.52 | 590 | 21.57 |
| **Gender** |  |  |  |  |
| Male | 1515 | 44.29 | 1157 | 42.3 |
| Female | 1810 | 52.91 | 1500 | 54.84 |
| Non-binary/Transgender | 96 | 2.81 | 78 | 2.85 |
| **Race/Ethnicity** |  |  |  |  |
| Non-Hispanic White | 2306 | 67.41 | 1881 | 68.78 |
| Hispanic | 500 | 14.62 | 364 | 13.31 |
| Non-Hispanic Black | 261 | 7.63 | 202 | 7.39 |
| Asian/PI | 222 | 6.49 | 186 | 6.8 |
| Other | 132 | 3.86 | 102 | 3.73 |
| **Education at baseline** |  |  |  |  |
| Less than high school | 37 | 1.08 | 29 | 1.06 |
| High school graduate | 278 | 8.13 | 206 | 7.53 |
| Some college | 815 | 23.82 | 640 | 23.4 |
| College graduate | 2291 | 66.97 | 1860 | 68.01 |
| **Household income** |  |  |  |  |
| Less than \$35,000 | 891 | 26.05 | 696 | 25.45 |
| \$35-49,999 | 392 | 11.46 | 309 | 11.3 |
| \$50-69,999 | 511 | 14.94 | 427 | 15.61 |
| \$70-99,999 | 609 | 17.8 | 495 | 18.1 |
| \$100,000+ | 1018 | 29.76 | 808 | 29.54 |
| **Employment at baseline** |  |  |  |  |
| Employed | 2175 | 63.58 | 1850 | 67.64 |
| Out of work | 378 | 11.05 | 179 | 6.54 |
| Homemaker | 165 | 4.82 | 131 | 4.79 |
| Student | 192 | 5.61 | 123 | 4.5 |
| Retired | 511 | 14.94 | 452 | 16.53 |
| **Setting** |  |  |  |  |
| Urban | 1460 | 42.68 | 1117 | 40.84 |
| Suburban | 901 | 26.34 | 752 | 27.5 |
| Rural | 1052 | 30.75 | 863 | 31.55 |
| Town | 8 | 0.23 | 3 | 0.11 |
| **Geographic region** |  |  |  |  |
| Northeast | 963 | 28.15 | 776 | 28.37 |
| Midwest | 632 | 18.47 | 485 | 17.73 |
| South | 960 | 28.06 | 796 | 29.1 |
| West | 862 | 25.2 | 674 | 24.64 |
| US Territories | 4 | 0.12 | 4 | 0.15 |
| **Healthcare worker** |  |  |  |  |
| No | 3073 | 89.83 | 2465 | 90.13 |
| Yes | 315 | 9.21 | 245 | 8.96 |
| Don't know | 33 | 0.96 | 25 | 0.91 |
| **Essential worker** |  |  |  |  |
| No | 2811 | 82.17 | 2272 | 83.07 |
| Yes | 610 | 17.83 | 463 | 16.93 |
| **Higher risk for severe COVID (V1 or V9)** |  |  |  |  |
| No | 2627 | 76.79 | 2045 | 74.77 |
| Yes | 794 | 23.21 | 690 | 25.23 |
| **Vaccination status at start of follow-up\*** |  |  |  |  |
| Un/under-vaccinated | 3421 | 100.00 | 2447 | 89.47 |
| Primary series only | 0 | 0.00 | 282 | 10.31 |
| Missing | 0 | 0.00 | 6 | 0.22 |
| **Vaccination status at end of follow-up\*** |  |  |  |  |
| Un/under-vaccinated | 3376 | 98.68 | 238 | 8.70 |
| Primary series | 18 | 0.53 | 251 | 9.18 |
| Boosted once | 0 | 0.00 | 1523 | 55.69 |
| 2+ boosters | 0 | 0.00 | 723 | 26.44 |
| Missing | 27 | 0.79 | 0 | 0.00 |

\* Serology period was defined as time from S1-S2 and S2-S3 for pre-vaccine and vaccine eras

*Vaccination status.* In the pre-vaccine/wild-type era, none of the 3,421 seronegative participants were vaccinated at the start of the follow-up (March 28, 2020), and only 18 participants (0.53%) had any vaccine doses as of November 17, 2020 (Table 1). In the vaccine/variant era, 282 (10.3%) of the 2,735 seronegative participants had completed a primary vaccine series as of February 10, 2021 (V6 questionnaire in Figure 1); 2,497 (91.3%) were fully vaccinated by the end of follow-up, including 2,246 (82.1%) boosted at least once and 723 (26.4%) boosted twice. In terms of the timing of vaccination, 87% percent of the vaccine/variant-era cohort had completed their primary series within 6 months of the start of follow-up in the vaccine/variant era (i.e., within 6 months of their seronegative specimen).

### Seroincidence of SARS-CoV-2 Infection

The seroincidence rate of SARS-CoV-2 infection in the vaccine/variant era cohort was nearly three times higher than in the pre-vaccine/wild-type era cohort. Specifically, we observed a SARS-CoV-2 infection rate of 9.61 per 100 person-years (95% CI 8.3-11.1) and 25.74 per 100 person-years (95% CI 24.2-27.3) in the pre-vaccine/wild-type era cohort and vaccine/variant era cohort, respectively (Table 2).

**Table 2.**
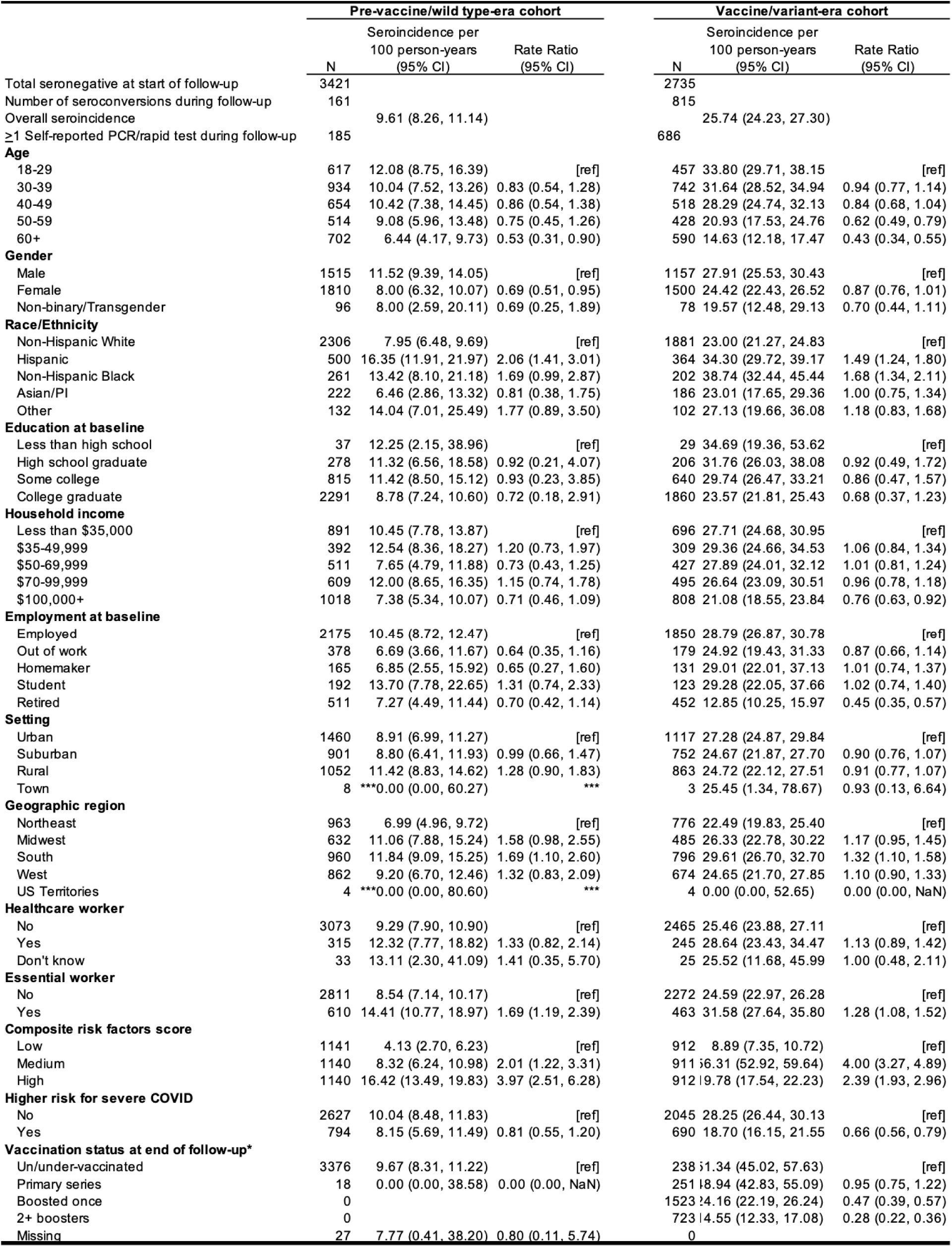
Crude seroincidence estimates in the pre-vaccine era and vaccine-era cohorts by sociodemographic factors and vaccination status.

#### Sociodemographic factors

Table 2 also shows the SARS-CoV-2 incidence rate and crude SARS-CoV-2 incidence rate ratios by sociodemographic factors and cohort era. Across the two cohorts, crude incidence rates were substantially higher in all sociodemographic subgroups in the vaccine/variant era cohort compared with the pre-vaccine/wild-type era cohort. Within each of the two cohorts, the SARS-CoV-2 infection rate varied substantially by sociodemographic factors, with *lower* SARS-CoV-2 infection in those aged 60 and older compared with 18-29 year-olds in both cohorts (IRR_pre-vaccine/wild-type_, 0.53 [95% CI, 0.31-0.90]; IRR_vaccine/variant_, 0.43 [95% CI, 0.34 -0.55]) and in women compared to men in the pre-vaccine/wild-type era cohort (IRR_pre-vaccine/wild-type_, 0.69 [95% CI, 0.51-0.95]). Some associations appeared to be protective only in the vaccine/variant era cohort (household income above $100,000 vs. less than $35,000, retired vs. employed, and higher vs. lower risk of severe COVID). In each cohort, we observed *higher* seroincidence of SARS-CoV-2 infection among Hispanic (IRR_pre-vaccine/wild-type_, 2.06 [95% CI, 1.41-3.01]; IRR_vaccine/variant_, 1.49 [95% CI: 1.24-1.80]) and non-Hispanic Black (IRR_pre-vaccine/wild-type_, 1.69 [95% CI, 0.99-2.87] ; IRR_vaccine/variant_, 1.68 [95% CI, 1.34-2.11]) participants compared with non-Hispanic White participants, essential workers compared with non-essential workers (IRR_pre-vaccine/wild-type_, 1.69 [95% CI, 1.19-2.39]; IRR_vaccine/variant_, 1.28 [95% CI, 1.08-1.52]), and in the South compared with the Northeast (IRR_pre-vaccine/wild-type_, 1.69 [95% CI, 1.10-2.60]; IRR_vaccine/variant_, 1.32 [95% CI 1.10-1.58]). However, some of these associations (Hispanic vs. non-Hispanic Whites, living in the Southern United States vs. the Northeast), became less pronounced in the vaccine/variant era cohort. Individuals who were at high risk for severe COVID-19 (vs. lower riks) also had significantly lower SARS-CoV-2 infection risk in the vaccine/variant era cohort (IRR_vaccine/variant_, 0.66 [95% CI: 0.56-0.79]), but not in the pre-vaccine/wild-type era cohort.

#### Vaccination status

In the vaccine/variant era cohort, the highest crude rates of SARS-CoV-2 infection were in un/under-vaccinated (51.34, 95% CI: 45.02-57.63) and fully vaccinated but not boosted participants (48.94, 95% CI: 42.83–55.09) (Table 2). Those who were fully vaccinated and received a booster had substantially lower rates of SARS-CoV-2 infection than the un/under-vaccinated, including those with one booster (IRR_vaccine/variant_, 0.47 [95% CI, 0.39-0.57]) and two or more boosters (IRR_vaccine/variant_, 0.28 [95% CI, 0.22-0.36]).

#### Epidemiologic risk factors

Table 3 shows the SARS-CoV-2 infection and crude SARS-CoV-2 infection rate ratios by epidemiologic risk factors that were present prior to or between serologic tests for each cohort. Crude incidence rates were substantially higher in most subgroups of epidemiologic risk factors in the vaccine/variant era cohort compared with the pre-vaccine/wild-type era cohort. In both cohorts, never social distancing with people you do not know (IRR_pre-vaccine/wild-type_, 3.16 [95% CI 1.47-6.77; IRR_vaccine/variant_, 2.00 [95% CI, 1.62-2.48] and never masking or sometimes masking in a variety of settings were associated with a higher risk of SARS-CoV-2 infection. Binge drinking (IRR_pre-vaccine/wild-type_, 1.47 [95% CI: 1.07-2.03]; IRR_vaccine/variant_, 1.45 [95% CI: 1.26-1.67]) and recent air travel (IRR_pre-vaccine/wild-type_, 1.49 [95% CI, 1.04-2.13]; IRR_vaccine/variant_, 1.20 [95% CI: 1.03-1.39]) were associated with higher risk of SARS-CoV-2 infection. A higher composite measure of risk was significantly associated with a higher risk of incident infection in both eras (Table 2).

**Table 3.** Crude seroincidence estimates in the pre-vaccine era and vaccine-era cohorts by epidemiologoc risk factors.

|  | Pre-vaccine/wild type-era cohort |  |  | Vaccine/variant-era cohort |  |  |
| --- | --- | --- | --- | --- | --- | --- |
|  | N | Seroincidence per 100 person-years (95% CI) | Rate Ratio (95% CI) | N | Seroincidence per 100 person-years (95% CI) | Rate Ratio (95% CI) |
| **COVID-19 risk factors** |  |  |  |  |  |  |
| Total seronegative at start of follow-up | 3421 |  |  | 2735 |  |  |
| Number of seroconversions during follow-up | 161 |  |  | 815 |  |  |
| Overall seroincidence |  | 9.61 (8.26, 11.14) |  |  | 25.74 (24.23, 27.30) |  |
| **Household characteristics** |  |  |  |  |  |  |
| **Household crowding** |  |  |  |  |  |  |
| Single-family with <4 members | 1373 | 8.50 (6.55, 10.94) | [ref] | 1129 | 20.53 (18.42, 22.81) | [ref] |
| Single-family with 4+ members | 625 | 9.20 (6.26, 13.24) | 1.08 (0.68, 1.71) | 494 | 32.69 (28.80, 36.82) | 1.59 (1.32, 1.92) |
| Multi-family with <4 members | 1160 | 9.18 (7.03, 11.88) | 1.08 (0.74, 1.57) | 917 | 28.49 (25.79, 31.34) | 1.39 (1.18, 1.63) |
| Multi-family with 4+ members | 171 | 17.58 (10.28, 28.08) | 2.07 (1.15, 3.71) | 128 | 33.67 (26.11, 42.12) | 1.64 (1.21, 2.23) |
| Dorms/Group homes/Other congregate settings | 17 | 11.56 (0.61, 50.66) | 1.36 (0.19, 9.82) | 13 | 18.33 (4.86, 45.50) | 0.89 (0.29, 2.79) |
| Other | 72 | 20.56 (9.33, 38.37) | 2.42 (1.10, 5.30) | 51 | 14.20 (7.09, 25.76) | 0.69 (0.36, 1.34) |
| **Children in the household** |  |  |  |  |  |  |
| No children in household | 2456 | 9.45 (7.90, 11.26) | [ref] | 2037 | 23.36 (21.69, 25.12) | [ref] |
| Children in household | 965 | 10.05 (7.50, 13.30) | 1.06 (0.75, 1.50) | 698 | 33.03 (29.75, 36.47) | 1.41 (1.22, 1.64) |
| **Household exposures** |  |  |  |  |  |  |
| No confirmed case in household member<sup>a</sup> | 3377 | 0.67 (0.57, 0.80) | [ref] | 2384 | 1.51 (1.39, 1.65) | [ref] |
| Confirmed case in household member<sup>a</sup> | 44 | 15.02 (10.30, 21.28) | 22.34 (14.77, 33.77) | 351 | 12.65 (11.29, 14.15) | 8.35 (7.22, 9.66) |
| **Social Distancing** |  |  |  |  |  |  |
| **Social distancing with people you know** |  |  |  |  |  |  |
| Always | 1136 | 9.26 (7.03, 12.07) | [ref] | 93 | 26.03 (18.39, 35.36) | [ref] |
| Sometimes | 1787 | 8.94 (7.18, 11.06) | 0.97 (0.68, 1.37) | 839 | 18.14 (15.83, 20.70) | 0.70 (0.47, 1.03) |
| Never | 300 | 16.74 (11.22, 24.09) | 1.81 (1.11, 2.94) | 1731 | 29.27 (27.28, 31.35) | 1.12 (0.77, 1.63) |
| NA | 158 | 5.31 (1.71, 13.74) | 0.57 (0.21, 1.59) | 72 | 33.39 (23.35, 45.08) | 1.28 (0.76, 2.18) |
| **Social distancing with people you do not know** |  |  |  |  |  |  |
| Always | 2615 | 8.85 (7.38, 10.58) | [ref] | 646 | 19.32 (16.64, 22.31) | [ref] |
| Sometimes | 660 | 10.39 (7.40, 14.35) | 1.17 (0.80, 1.72) | 1468 | 24.07 (22.08, 26.18) | 1.25 (1.03, 1.50) |
| Never | 53 | 27.94 (12.85, 49.52) | 3.16 (1.47, 6.77) | 466 | 38.66 (34.39, 43.10) | 2.00 (1.62, 2.48) |
| NA | 53 | 15.99 (5.25, 36.89) | 1.81 (0.67, 4.90) | 155 | 33.58 (26.75, 41.14) | 1.74 (1.29, 2.35) |
| **Spent time in public places** |  |  |  |  |  |  |
| Did not attend mass gatherings | 3071 | 9.55 (8.13, 11.18) | [ref] | 2460 | 24.97 (23.41, 26.61) | [ref] |
| Attended mass gatherings | 350 | 10.08 (6.25, 15.70) | 1.06 (0.65, 1.72) | 275 | 32.80 (27.64, 38.40) | 1.31 (1.07, 1.62) |
| **Indoor dining/bar** |  |  |  |  |  |  |
| No indoor dining/bar | 1666 | 6.52 (4.96, 8.50) | [ref] | 391 | 20.12 (16.67, 24.06) | [ref] |
| Indoor dining/bar | 1755 | 12.52 (10.42, 14.96) | 1.92 (1.38, 2.67) | 2344 | 26.73 (25.08, 28.46) | 1.33 (1.07, 1.64) |
| **Outdoor dining/bar** |  |  |  |  |  |  |
| No outdoor dining/bar | 1551 | 8.75 (6.85, 11.09) | [ref] | 674 | 24.48 (21.57, 27.64) | [ref] |
| Outdoor dining/bar | 1870 | 10.28 (8.45, 12.43) | 1.17 (0.86, 1.61) | 2061 | 26.16 (24.41, 27.99) | 1.07 (0.91, 1.25) |
| **Place of worship** |  |  |  |  |  |  |
| Did not visit place of worship | 3062 | 8.78 (7.42, 10.35) | [ref] | 2017 | 23.83 (22.13, 25.61) | [ref] |
| Visited place of worship | 359 | 16.81 (11.72, 23.42) | 1.91 (1.28, 2.86) | 718 | 31.38 (28.20, 34.75) | 1.32 (1.14, 1.53) |
| **Public park/pool** |  |  |  |  |  |  |
| Did not visit public park/pool | 1087 | 9.48 (7.14, 12.46) | [ref] | 529 | 23.80 (20.57, 27.34) | [ref] |
| Visited public park/pool | 2334 | 9.66 (8.06, 11.53) | 1.02 (0.73, 1.43) | 2206 | 26.22 (24.53, 27.99) | 1.10 (0.92, 1.31) |
| **Gathered in groups >=10** |  |  |  |  |  |  |
| No | 2392 | 8.65 (7.13, 10.44) | [ref] | 422 | 19.87 (16.57, 23.63) | [ref] |
| Indoors only | 195 | 18.20 (11.25, 27.82) | 2.10 (1.26, 3.52) | 307 | 25.67 (21.29, 30.57) | 1.29 (0.97, 1.71) |
| Outdoors only | 453 | 9.27 (5.97, 14.02) | 1.07 (0.67, 1.72) | 231 | 23.14 (18.32, 28.74) | 1.16 (0.85, 1.60) |
| Indoors and outdoors | 353 | 12.11 (7.83, 18.14) | 1.40 (0.88, 2.24) | 1775 | 27.60 (25.67, 29.61) | 1.39 (1.13, 1.71) |
| **Mask use** |  |  |  |  |  |  |
| **Mask while grocery shopping** |  |  |  |  |  |  |
| Always | 3083 | 9.24 (7.85, 10.84) | [ref] | 1461 | 19.53 (17.70, 21.50) | [ref] |
| Sometimes | 132 | 20.31 (11.37, 33.16) | 2.20 (1.22, 3.96) | 693 | 29.65 (26.50, 33.00) | 1.52 (1.28, 1.79) |
| Never | 33 | 99.00 (93.76, 99.95) | 0.00 (0.00, NaN) | 408 | 40.89 (36.35, 45.59) | 2.09 (1.75, 2.50) |
| Did not go grocery shopping | 133 | 9.26 (3.82, 19.72) | 1.00 (0.44, 2.27) | 173 | 29.79 (23.62, 36.76) | 1.53 (1.16, 2.01) |
| **Mask while indoors visiting non-household members** |  |  |  |  |  |  |
| Always | 1118 | 7.76 (5.74, 10.39) | [ref] | 154 | 16.89 (12.00, 23.16) | [ref] |
| Sometimes | 1131 | 11.30 (8.85, 14.29) | 1.46 (0.99, 2.14) | 761 | 19.78 (17.24, 22.59) | 1.17 (0.80, 1.71) |
| Never | 463 | 15.12 (10.78, 20.73) | 1.95 (1.24, 3.07) | 1701 | 29.64 (27.62, 31.73) | 1.75 (1.23, 2.50) |
| NA (did not visit indoors) | 668 | 5.88 (3.67, 9.18) | 0.76 (0.44, 1.30) | 119 | 21.32 (15.05, 29.18) | 1.26 (0.77, 2.08) |
| **Mask while indoors at work** |  |  |  |  |  |  |
| Always | 1372 | 10.53 (8.36, 13.15) | [ref] | 811 | 21.71 (19.16, 24.48) | [ref] |
| Sometimes | 299 | 12.21 (7.60, 18.87) | 1.16 (0.69, 1.95) | 747 | 30.49 (27.40, 33.75) | 1.40 (1.17, 1.69) |
| Never | 68 | 19.40 (8.14, 38.13) | 1.84 (0.80, 4.24) | 380 | 38.50 (33.90, 43.31) | 1.77 (1.45, 2.18) |
| NA (did not attend indoor workplace) | 1641 | 7.87 (6.14, 10.01) | 0.75 (0.53, 1.05) | 797 | 19.82 (17.35, 22.54) | 0.91 (0.75, 1.11) |
| **Mask while at salon/gym** |  |  |  |  |  |  |
| Always | 1527 | 9.90 (7.91, 12.31) | [ref] | 982 | 19.22 (17.02, 21.62) | [ref] |
| Sometimes | 180 | 23.10 (15.00, 33.63) | 2.33 (1.42, 3.82) | 656 | 28.49 (25.29, 31.91) | 1.48 (1.23, 1.79) |
| Never | 108 | 13.91 (6.24, 27.20) | 1.40 (0.65, 3.05) | 645 | 35.97 (32.47, 39.62) | 1.87 (1.57, 2.24) |
| NA (did not attend) | 1566 | 7.37 (5.66, 9.53) | 0.74 (0.53, 1.05) | 452 | 22.48 (19.07, 26.29) | 1.17 (0.94, 1.46) |
| **Mask while on public transit** |  |  |  |  |  |  |
| Always | 758 | 10.32 (7.52, 13.94) | [ref] | 1123 | 24.08 (21.80, 26.52) | [ref] |
| Sometimes | 32 | 28.82 (9.67, 58.36) | 2.79 (1.00, 7.82) | 109 | 42.93 (33.99, 52.34) | 1.78 (1.33, 2.40) |
| Never | 80 | 13.71 (5.16, 29.94) | 1.33 (0.52, 3.37) | 268 | 39.32 (33.75, 45.17) | 1.63 (1.32, 2.02) |
| NA (did not use) | 2510 | 8.98 (7.47, 10.76) | 0.87 (0.60, 1.25) | 1235 | 23.05 (20.92, 25.32) | 0.96 (0.82, 1.12) |
| **Outdoor mask use** |  |  |  |  |  |  |
| No mask use outdoors | 707 | 11.04 (7.98, 15.01) | [ref] | 182 | 32.40 (26.13, 39.36) | [ref] |
| Mask use outdoors | 2714 | 9.25 (7.78, 10.96) | 0.84 (0.58, 1.21) | 2553 | 25.28 (23.73, 26.89) | 0.78 (0.61, 1.00) |
| **Movement during the pandemic** |  |  |  |  |  |  |
| **Use of public transit** |  |  |  |  |  |  |
| Avoided or did not use | 730 | 10.29 (7.44, 14.02) | [ref] | 601 | 28.19 (24.87, 31.76) | [ref] |
| Used public transit | 2691 | 9.42 (7.92, 11.16) | 0.92 (0.63, 1.32) | 2134 | 25.06 (23.38, 26.83) | 0.89 (0.76, 1.05) |
| **Recent air travel** |  |  |  |  |  |  |
| No | 2839 | 8.84 (7.42, 10.50) | [ref] | 2019 | 24.51 (22.80, 26.31) | [ref] |
| Yes | 582 | 13.16 (9.63, 17.67) | 1.49 (1.04, 2.13) | 716 | 29.34 (26.23, 32.64) | 1.20 (1.03, 1.39) |
| **Alcohol/substance use** |  |  |  |  |  |  |
| **Binge drinking** |  |  |  |  |  |  |
| No | 2470 | 8.50 (7.02, 10.25) | [ref] | 1878 | 22.64 (20.92, 24.46) | [ref] |
| Yes | 951 | 12.51 (9.71, 15.95) | 1.47 (1.07, 2.03) | 857 | 32.79 (29.85, 35.86) | 1.45 (1.26, 1.67) |
| **Regular cannabis or un-prescribed opioid use** |  |  |  |  |  |  |
| No | 2953 | 9.75 (8.29, 11.42) | [ref] | 2146 | 25.26 (23.58, 27.03) | [ref] |
| Yes | 468 | 8.71 (5.54, 13.32) | 0.89 (0.56, 1.43) | 589 | 27.46 (24.17, 31.01) | 1.09 (0.92, 1.28) |

<sup>a</sup> Person-time is in months

#### Changes in epidemiologic risk factors between pre-vaccine and vaccine/variant eras

Some new associations emerged that were not present in the pre-vaccine/wild-type era cohort (Table 3). In the pre-vaccine/wild-type era cohort, people living with <4 household members in a single unit of a multi-unit dwelling and those living with 4 or more household members in a single-family dwelling had similar incidence as those living in single-family dwellings with <4 household members. But in the vaccine/variant era cohort, those living with <4 household members in a single unit of a multi-unit dwelling and those living with 4 or more household members in a single-family dwelling saw their risk increase compared with those living in single-family dwellings with <4 household members (IRR_vaccine/variant_, 1.39 [95% CI, 1.18-1.63] for multi-unit dwelling with <4 household members; IRR_vaccine/variant_, 1.59 [95% CI: 1.32-1.92] for single-family dwelling with 4+ household members). Similarly, as shown in Table 3, having a child in the household was not associated with a higher risk of SARS-CoV-2 infection in the pre-vaccine/wild-type era cohort, but was in the vaccine/variant-era cohort (IRR_vaccine/variant_, 1.41 [95% CI, 1.22-1.64]). Social distancing ‘with people you don’t know’ sometimes (vs. always) was not associated with a higher risk of SARS-CoV-2 infection in the pre-vaccine/wild-type era cohort, but was significantly associated with a higher risk in the vaccine/variant era cohort (IRR_vaccine/variant_, 1.25 [95% CI, 1.03–1.50]). Associations for some risk factors persisted but became less pronounced in the vaccine/variant era cohort compared with the pre-vaccine/wild-type era cohort (Table 3). Specifically, having a confirmed case in the household had the highest absolute incidence rate and was the strongest risk factor in each cohort, but the strength of the association decreased in the vaccine/variant-era cohort (IRR_vaccine/variant_, 8.35 [95% CI, 7.22-9.66]) vs. the pre-vaccine/wild-type era cohort (IRR_pre-vaccine/wild-type_, 22.34 [95% CI, 14.77– 33.77]). Social distancing ‘with people you know’ never (vs. always) was associated with a higher risk of SARS-CoV-2 infection in the pre-vaccine/wild-type era cohort (IRR_pre-vaccine/wild-type_, 3.16 [95% CI, 1.47-6.77]) than in the vaccine/variant era cohort (IRR_vaccine/variant_, 2.00 [95% CI, 1.62-2.48]). The risk ratio for SARS-CoV-2 infection was also higher in the pre-vaccine/wild-type era cohort than the vaccine/variant era cohort for indoor dining, visiting a place of worship, gathering indoors with 10 or more persons, and social distancing with ‘people you do not know’ never (vs. always).

Associations of mask use with incidence depended on the context. For mask use while grocery shopping, at the salon or gym, or on public transit, risk for sometimes use (vs. always) was elevated in both cohorts but was less pronounced in the vaccine/variant era cohort (Table 3). No mask use (vs. always) while indoors visiting non-household members was associated with an elevated risk of infection in both cohorts, but less so in the vaccine/variant era cohort. Mask use sometimes or never (vs. always) while indoors at work was associated with a higher risk in the vaccine/variant era, but not in the pre-vaccine/wild-type era, as was no mask use (vs. always) while at the salon or gym or while on public transit.

### Poisson models of SARS-CoV-2 seroconversion in strata of risk factors

Table 4 shows adjusted IRRs and 95% CIs from the overall multivariable model and the 12 risk-factor group-specific multivariate Poisson models stratified by vaccine status in the vaccine/variant era, with the pre-vaccine/wild-type cohort as the referent group, adjusting for age, gender, and presence of co-morbidities. Relative to the pre-vaccine/wild-type cohort, the risk of SARS-CoV-2 infection was similar between un/undervaccinated and fully vaccinated participants in the vaccine/variant era. However, the risk of infection tended to decrease with an increasing number of boosters. Specifically, adjusted incident rate ratios (aIRR) with the pre-vaccine/wild-type cohort as the referent group were: aIRR_un/undervaccinated_=5.3 (95% CI 4.2-6.7); aIRR_primary series only_=5.1 (95%CI: 4.1-6.4); aIRR_boosted once_=2.5 (95% CI 2.1-3.0), and aIRR_boosted twice_=1.65 (95%CI: 1.3-2.1) (Table 4; Figure 3). These associations were essentially unchanged within risk factor-stratified models, except for those with a confirmed case of SARS-CoV-2 in the household, where the relative change in SARS-CoV-2 infection between the pre-vaccine/wild-type cohort and the vaccine/variant-era cohort was smaller than that in other risk groups.

**Figure 3.**
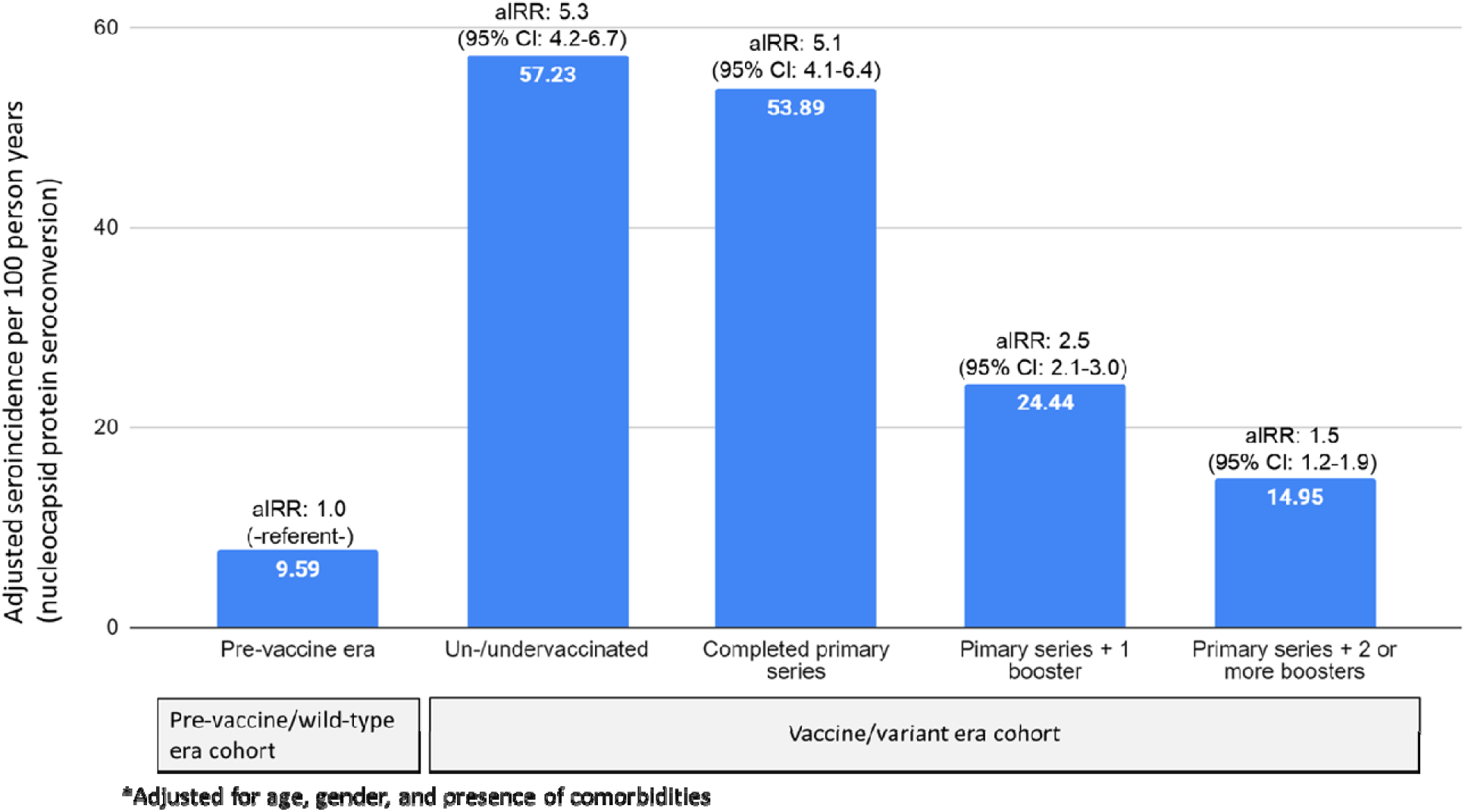
Adjusted* incidence rates and incidence rate ratios (aIRR) for SARS-CoV-2 infection by vaccination status compared with the pre-vaccine era

**Table 4.**
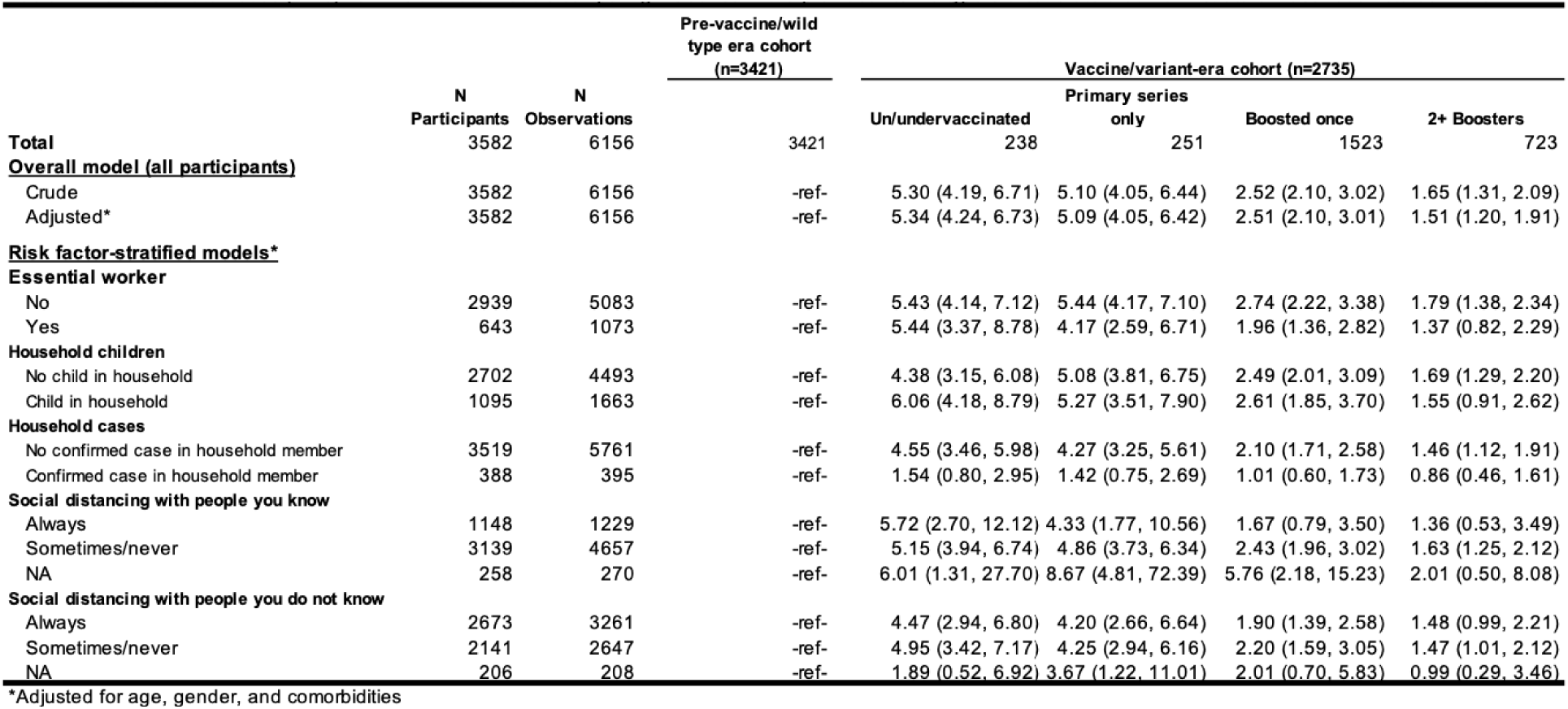
Incidence Rate Ratios (IRRs) from multivariate models comparing incidence in the pre-vaccine/wild-type era cohort to that within strata of vaccination status in the.

### Self-reported testing

Table 5 shows the number and proportion of participants who tested positive on serologic tests as part of our study as well as on self-reported PCR or rapid tests taken by participants outside of the study for each cohort. Compared to serologic test results, participants had lower rates of test positivity on self-reported viral PCR or rapid tests. Specifically, in the pre-vaccine/wild-type era cohort, 4.0% (n=137) of participants self-reported a positive PCR or rapid test outside of the study, compared to 4.7% (n=161) that tested positive on serologic testing (ratio 85%). In the vaccine/variant era cohort, 21% (n=561) self-reported a positive test, compared to 30% (n=815) from serologic testing (ratio 69%). Thus, the proportion of participants with an infection detected outside the study declined from 85% in the pre-vaccine/wild-type era cohort to 69% in the vaccine/variant era cohort (Table 5).

**Table 5.** Positivity rate of serologic testing compared with self-reported PCR/rapid testing.

|  | (n=3,421) |  | (n=2,735) |  |
| --- | --- | --- | --- | --- |
|  | Number positive | Positivity rate | Number positive | Positivity rate |
| Positive serologic test (study) | 161 | 4.71% | 815 | 29.80% |
| Self-reported positive PCR/rapid test (outside study) | 137 | 4.00% | 561 | 20.51% |
| Ratio of self report/serology positivity rates | 85% |  | 69% |  |

## DISCUSSION

In a community-based prospective study with repeat serologic testing of SARS-CoV-2 n protein seronegative individuals, we observed a nearly 3-fold increase in the incidence of n protein seroconversion, coinciding with the SARS-CoV-2 vaccine/variant era (25.74 per 100 person-years) as compared with the pre-vaccine/wild-type era (9.61 per 100 person-years). This corresponds to an increase in SARS-CoV-2 infection risk from 10% to 26% of participants infected per year. The large increase in SARS-CoV-2 incidence coincided with a relaxing of guidelines (e.g., around social distancing, masking, school attendance, in-person school attendance) and with surges of increasingly transmissible, immune evasive variants: Alpha (March–June 2021)^46^, Delta (June–December 2021) and Omicron variant and subvariants (December 2021-present)^47^, all emerged as vaccines were being more widely taken up. Our cohort findings are consistent with widespread community transmission in the general population, particularly during the Delta and Omicron surges, including in workplaces and households with children in the vaccine/variant era compared with the pre-vaccine/wild-type era.^48–52^ Despite the large increase in community transmission in the vaccine/variant era, being more up-to-date on vaccines (i.e., being boosted) was associated with a lower risk of SARS-CoV-2 infection compared with being un/undervaccinated or only receiving the primary vaccine series. While there are likely differences in people who were boosted compared with those who were not, these associations were maintained across several epidemiologic risk strata. Although being boosted was associated with a reduced incidence in the vaccine/variant era, except for the groups reporting a confirmed case in the household, the incidence was still generally 1.3-2 times higher among individuals with 2+ boosters compared with those in the pre-vaccine/wild-type era cohort (Table 4). While this highlights the potential for new variants to cause breakthrough infections even among those who are more up-to-date on vaccines, it also suggests that being up-to-date on SARS-CoV-2 vaccines can greatly reduce the risk of infection during surges. In addition, many non-pharmaceutical interventions used by cohort participants (e.g., masking in many different settings, social distancing) remained associated with substantially lower SARS-CoV-2 incidence rates in the vaccine/variant-era cohort, despite large increases in absolute incidence rates (Table 3).

In multivariate models, those who only received the primary vaccine series and no booster doses had similar SARS-CoV-2 infection risk as those who were un/undervaccinated in the vaccine/variant era. In both groups, the risk was approximately 5 times higher than in the pre-vaccine/wild-type era cohort. However, receipt of booster doses beyond the primary vaccine series was associated with a lower risk of infection compared with other vaccine status groups in the vaccine/variant era cohort. In fact, the risk of infection became progressively lower as the number of vaccine booster doses increased. This could be because 87% of the vaccine/variant-era cohort was fully vaccinated by August 2021 (∼six months into cohort follow-up, Table 1), and enough time had passed such that boosters would have been needed for most participants in order to offer some protection against infection from the more immune evasive variants.

There also may be differences in behaviors and other factors among these groups. Importantly, however, these associations were observed in each stratum across various risk factors, with models that adjusted for age, gender, and presence of comorbidities.

Our study showed substantial increases in SARS-CoV-2 incidence rates in the vaccine/variant era cohort compared to the pre-vaccine/wild-type era cohort within every sociodemographic subgroup and epidemiologic risk factor that we examined. Importantly, many factors that appeared protective against SARS-CoV-2 in the pre-vaccine/wild-type era cohort (e.g., NPIs such as masking and social distancing) remained protective in the vaccine/variant era cohort, despite the major increases in community transmission that occurred. This suggests that NPIs may play an important role in limiting SARS-CoV-2 transmission even during major surges of new variants, and in protecting those most vulnerable to SARS-CoV-2 infection.

However, it should be noted that while the incidence rates in those engaging in protective behaviors in the vaccine/variant era cohort were lower than those who didn’t engage in protective behaviors, absolute incidence rates were still very high in most instances in the vaccine/variant era cohort compared with the pre-vaccine/wild-type era cohort, highlighting that protective behaviors can reduce *but not eliminate* risk when community transmission rates are high. For example, there was a lower infection risk associated with wearing masks at work in the vaccine/variant era cohort, but the absolute incidence rate in those wearing masks at work was also much higher than in the pre-vaccine/wild-type era cohort. This could be due to higher levels of exposure inside the workplace, more individuals returning to in-person work, or higher levels of exposure from other sources (such as at home or on public transportation) or other locations that may not have been open during the pre-vaccine/wild-type era (e.g., gyms, indoor dining).

We also noted some new epidemiologic risk factors that emerged in the vaccine/variant cohort (e.g., having a child in the household), likely reflecting the higher incidence of infection in the general population, including new sources of exposure such as children attending in-person school. Lastly, looking at the composite risk score, there was a clear dose-response in the pre-vaccine/wild-type era cohort, but in the vaccine/variant era, the highest level of the composite risk score was lower than that of the medium risk score. This could be because the risk associated with having a case in the household, by far the strongest risk factor in both eras (Table 3), cannot increase as much in comparison with that of other risk factors when moving from the pre-vaccine/wild-type era to the vaccine/variant era (i.e., a ceiling effect).

Our study aligns with recent cross-sectional, population-representative surveys that were conducted during Omicron variant surges in some ways and not others. For example, similar to our study, recent NYC-based and national surveys conducted during major surges found high absolute point prevalence of SARS-CoV-2 infection but substantially lower relative point prevalence estimates among older (vs. younger) adults, those with comorbidities (vs. those without), and higher relative point prevalence estimates among those in households with school-aged children (vs. those without).^53,54^ However, in contrast to our study, these surveys found that vaccinated and boosted respondents had similar point prevalence estimates of SARS-CoV-2 infection to unvaccinated respondents. Reasons for this discrepancy could be that the surveys captured self-reported infection (positive point of care test, home test, or symptoms plus close contact) during the two weeks prior to the survey, while our study examined SARS-CoV-2 infection prospectively, as measured by serology over a longer time frame. In our cohort, compared with the seropositivity rate, the positivity rate on self-reported PCR/rapid tests over the same time period was lower in both the pre-vaccine/wild-type (85%) and vaccine/variant era cohorts (69%; Table 5). The reasons for the lower ratio in the vaccine/variant era cohort are not clear, but may be due to the fact that this was a highly vaccinated cohort, and fully vaccinated individuals with breakthrough infections may be less symptomatic, have a lower viral load, and experience a shorter duration of infection/illness.^55–57^ As such, these participants may not have recognized signs of SARS-CoV-2 infection and/or felt the need to test for SARS-CoV-2.

As the COVID-19 pandemic evolves, it remains important to monitor the incidence of SARS-CoV-2 infections. It may become more difficult, however, to identify cases through routine provider/laboratory reporting of PCR or rapid antigen tests. Individuals are increasingly less likely to be required to test in certain scenarios, may choose not to test, or may exclusively use at-home tests that are not captured in routine surveillance. Thus, using serologic testing in cohort studies is a useful strategy to characterize SARS-CoV-2 incidence and risk factors.

Strengths of our study include its prospective nature, with time-updated exposure measurement prior to outcome ascertainment. We also used repeat serologic testing to examine SARS-CoV-2 nucleocapsid seroconversion as the outcome measure of incident infection. Our design, which compared incidence in the two cohorts, leveraged the use of individuals as their own controls, since 75% of the overall sample was represented in both cohorts (Figure 1), helping to reduce confounding when comparing incidence in the two eras. Finally, comparing incidence rates within models specific to different strata of risk factors also helps to limit confounding of the association of vaccination status with incidence by risk behaviors.

Our study also has limitations worth noting. The observed cumulative incidence in our cohort may be lower than the true cumulative incidence in our cohort because of the imperfect nature of serologic testing and the potential waning of SARS-CoV-2 antibodies,^58^ particularly for milder infections.^59,60^ Studies of SARS-CoV-2 antibody persistence have suggested the waning of antibodies to both nucleocapsid and spike proteins.^61,62^ Boosted individuals who have an infection after vaccination may experience a more rapid waning of nucleocapsid antibodies.^63^ Our study required total nucleocapsid seronegativity for inclusion. Because of the timing of specimen collection relative to infection in our cohort (median of 191 days in the pre-vaccine/wild type era cohort and 476 days in the vaccine/variant era cohort), this could mean that we have underestimated the true cumulative incidence due to waning. Additionally, immunocompromised status for study participants was not collected, and fully vaccinated status could therefore not be accurately assigned using three doses among this subgroup. For these participants, the third primary series dose may have been misidentified as a booster dose or skipped entirely, resulting in a fully vaccinated status. We did, however, adjust for the presence of comorbidities.

Crude associations between SARS-CoV-2 risk factors and incidence are subject to confounding. For example, behaviors between risk groups likely differ, with interpretation for some associations further hampered by small sample sizes in some exposure strata. Some risk behaviors may have been underreported (e.g., due to social desirability), which would bias observed associations toward the null. While our study was prospective, because we used a midpoint method to infer the timing of infection between a negative and positive serologic test it is possible that some measured exposures, including vaccination, did not temporally precede infection. Also, some infections may have occurred in between vaccine doses. Finally, while our study was able to examine the role of behavioral risk factors over time, because of the timing of serologic testing, we could not distinguish the variant-specific effects (e.g., wild-type vs. Alpha, Delta vs. Omicron).

## Conclusion

Increases in the incidence of infection and newly emerging risk factors in the vaccine/variant era likely resulted from multiple co-occurring factors related to policy changes, individual- and community-level behavior changes (due to the availability of vaccines and relaxation of restrictions), and changing virus properties (i.e., more transmissible, immune evasive variants). While SARS-CoV-2 incidence increased markedly in most groups in the vaccine/variant cohort, being up to date on vaccines and the use of NPIs (masking, distancing) likely reduced the risk of SARS-CoV-2 infection during major surges, making them relevant strategies to mitigate the impact of future SARS-CoV-2 surges, including those due to new variants that may evade existing vaccine-induced and hybrid immunity. As the COVID-19 pandemic transitions to the endemic phase, serologic testing is a useful strategy to characterize SARS-CoV-2 incidence and risk factors.

## CONTRIBUTORS

DN, MMR, AS conceptualized the study. AS performed statistical analyses. DN and MSR wrote the first draft of the paper. All authors contributed to interpreting the data and revising of the manuscript. SGK, WY, MMR AB, SK, and DN contributed to data collection, cleaning and management. DN, MMR, SGK, and CG contributed to obtaining funding for the research.

## Data Availability

There is a public-use dataset for the CHASING COVID Cohort Study. Other data are available upon reasonable request from the authors.
DOI: 10.5281/zenodo.7305435

https://zenodo.org/record/7305435

## ACKNOWLEDGEMENTS

The authors wish to thank the participants of the CHASING COVID Cohort Study. We are grateful to you for your contributions to the advancement of science around the SARS-CoV-2 pandemic. We are grateful to MTL Labs for serologic testing of our cohort’s specimens.

## STATISTICAL APPENDIX

*LASSO.* The study samples of 3,421 and 2,735 participants for the pre-vaccine/wild type and post-vaccine variant eras respectively were split randomly into equally sized training and test data sets. A grouped LASSO regression was fit using training data and 10-fold cross validation was used to obtain the minimum value of lambda (the tuning parameter). The variables included in the LASSO model were household crowding, having children in the household, confirmed COVID-19 case in a household member, attending mass gatherings, indoor dining in a restaurant/bar, outdoor dining in a restaurant/bar, visiting places of worship, visiting public park or pool, gathering with groups of 10 or more indoors and/or outdoors, mask use while grocery shopping, mask use while visiting non-household members, mask use indoors in gym/salons, mask use indoors at work, mask use outdoors, using public transit, recent air travel, alcohol use, and substance use. Using the minimum lambda value, a grouped LASSO model was run on the entire dataset (training + test) to obtain factors that best predicted seroconversion. The LASSO model selected having a confirmed COVID-19 case in a household member, indoor dining in a bar/restaurant, gathering with groups of 10 or more outdoors, and mask use indoors in salons/gyms as the most predictive of seroconversion in our cohort. A logistic regression model was fitted using these variables selected by LASSO regression with seroconversion (Y/N) as the outcome. Coefficients of the variables were multiplied by 10 and rounded up to the nearest integer to create a score associated with that variable/risk factor. If a participant engaged in a given risk factor, they were assigned the score associated with that risk factor. Scores were then summed across risk factors, normalized between 0 and 100, and a total composite risk score was created for each participant.

